# Anti-mitochondrial antibodies in systemic sclerosis target enteric neurons and are associated with GI dysmotility

**DOI:** 10.1101/2024.11.26.24317983

**Authors:** Zsuzsanna H. McMahan, Livia Casciola-Rosen, Timothy Kaniecki, Laura Gutierrez-Alamillo, Su Hong Ming, Philippa Seika, Subhash Kulkarni

## Abstract

**Background:** Most patients with systemic sclerosis (SSc) experience gastrointestinal (GI) dysmotility. The enteric nervous system (ENS) regulates GI motility, and its dysfunction causes dysmotility. A subset of SSc patients harbor autoantibodies against the M2 mitochondrial antigen (AM_2_A). Here, we investigate whether M2 is expressed by specific ENS cells, and if AM_2_A associate with GI dysmotility in SSc patients.

**Methods:** Sera from 154 well-characterized patients with SSc were screened for AM_2_A by ELISA. Clinical features and GI transit data were compared between AM_2_A-positive and negative patients. HepG2 cells were cultured with these sera and co-stained with AM_2_A.

**Results:** Nineteen of 147 patients (12.9%) were AM_2_A positive. AM_2_A positivity was significantly associated with slower transit in the esophagus (β -14.4, 95%CI -26.2, -2.6) and stomach (β -7.9, 95% CI -14.1, -1.6). Immunostaining demonstrated pan-mitochondrial antigens TOM-20 and M2 enrichment in human ENS neurons, specifically in mesoderm-derived enteric neurons (MENS). HepG2 cells cultured with SSc sera showed that SSc autoantibodies penetrate live cells and that AM_2_A and other SSc autoantibodies are localized to subcellular compartments containing target antigens.

**Conclusion:** AM_2_A in SSc patients associate with slower GI transit. MENs are enriched in mitochondria, suggesting enhanced susceptibility to mitochondrial dysfunction and associated GI dysmotility in SSc. Our finding that SSc autoantibodies penetrate live cells *in vitro* suggests that SSc-AM_2_A may penetrate MENs *in vivo,* driving ENS and GI dysfunction. Further studies are warranted to understand whether AM_2_As contribute to mitochondrial dysfunction, and whether mitochondrial dysfunction contributes to GI dysmotility in SSc.

**Key messages:**

- What is already known on this topic

- subset of SSc patients have autoantibodies against the M2 mitochondrial antigen (AM_2_A). Whether AM_2_A antibodies inform the presence or severity of GI dysfunction in SSc is unknown.

- What this study adds:

- AM_2_A antibodies in SSc patients associate with slower upper GI transit.
- Mitochondria are enriched in a recently identified mesoderm-derived lineage of enteric neurons (MENs), which play a major role in GI motility. This suggests that MENS may be more mitochondrial-dependent than other cell types, and thus more susceptible to mitochondrial dysfunction. This may contribute to dysmotility in AM_2_A-positive SSc patients.
- SSc autoantibodies penetrate live cells *in vitro* and bind to their target antigens intracellularly.

- How this study might affect research, practice or policy

- AM_2_A antibodies in SSc patients may penetrate MENs in vivo, driving ENS dysfunction and subsequent GI dysmotility
- This potentially novel mechanism in SSc GI disease could inform our current approach to diagnosing and managing these patients.

## INTRODUCTION

Systemic sclerosis (SSc) is a chronic and progressive autoimmune disease characterized by vascular abnormalities and degenerative and fibrotic changes in the skin, joints, and internal organs. Of the various internal organs affected, gastrointestinal (GI) involvement is the most common^1^. GI dysfunction, which often manifests as loss of healthy GI tract motility, impacts more than 80% of SSc patients. Furthermore, the severity of GI dysfunction in SSc is significantly correlated with a worse prognosis^1–3^. We previously documented the extent and severity of GI dysfunction and dysmotility in SSc patients^3–12^. We have also identified novel intracellular autoantigens targeted in SSc patients with significant GI dysfunction (SSc-GI)^13^. The discovery of new autoantigens targeted in SSc patients suggests that a comprehensive identification of all targeted autoantigens associated with SSc-GI disease has not yet been achieved. This deficit precludes us from understanding the serological diversity in SSc-GI patients, and how this diversity informs the heterogeneous presentation of GI complications in SSc patients. Knowledge of these antibodies will also provide important insights into which biological pathways are relevant across SSc-GI clinical subsets.

Amongst the autoantibodies present in SSc patient sera, anti-mitochondrial antibodies (AMA), specifically targeting the mitochondrial M2 antigen, are found in 8 – 15% of patients^14,15^. Fregeau et al. determined that all AMA in limited SSc/CREST syndrome patients were immunoreactive against the M2 antigen^16^, which is designated as AM_2_A. Antibodies against AM_2_A are an important diagnostic marker for Primary Biliary Cholangitis (PBC), a condition in which patients also experience esophageal dysmotility^17^. Notably, although antibodies against AM_2_A are found in a significant subset of SSc patients, and are associated with GI dysfunction in PBC patients, the association between GI dysfunction and AM_2_A prevalence in patients with SSc remains unknown.

AMA (specifically anti-AM_2_A) are thought to arise either from primary disease-driven cellular damage that results in the exocytosis of mitochondrial antigens or to be disease-driving autoantibodies that impair mitochondrial function^18, 19^. In either scenario, mitochondrial dysfunction is associated with the presence of AMA. Mitochondrial disorders (MID), arising from genetic abnormalities in mitochondria, are associated with severe GI dysfunction. This often manifests as poor appetite, gastroesophageal reflux disease (GERD), dysphagia, vomiting, gastroparesis, constipation, GI pseudo-obstruction, and diarrhea^20, 21^. The presence of severe GI dysfunction in MID, whose manifestations mirror SSc GI dysfunction, and the presence of AMA in SSc patients raises questions about the prevalence of AMA/ AM_2_A in SSc patients with significant GI dysfunction and whether AM_2_A levels associate with GI severity in SSc patients.

This study assessed the AM_2_A status in a cohort of SSc patients with well-characterized GI dysfunction. We found that AM_2_A are present in ∼13% of patients with SSc and GI disease and that these antibodies are associated with slower GI transit. We also determined that mitochondria are enriched in a recently identified mesoderm-derived lineage of enteric neurons (MENs), which play a major role in GI motility. This suggests that MENS may be more mitochondrial-dependent and thus more susceptible to mitochondrial dysfunction compared to other cell types. Finally, using an immunofluorescence-based approach, we demonstrated that these antibodies are internalized, and navigate to and bind mitochondria intracellularly, providing an opportunity for AM_2_A to impact function. These findings provide intriguing new insights into the potential etiology of GI dysfunction in patients with SSc GI disease.

## METHODS

### Patient cohorts

The Gastrointestinal Assessment Protocol (GAP) Cohort is a prospectively enrolled National Institutes of Health (NIH)–sponsored GI cohort of well-characterized patients with SSc from the Johns Hopkins Scleroderma Center. It includes a subset of the Johns Hopkins Scleroderma Center Research Registry. Patients were consecutively recruited during routine clinical visits from July 2016 to April 2022 if they met either the 2013 American College of Rheumatology (ACR)/EULAR criteria for SSc or had at least three of the five features of CREST syndrome (calcinosis, Raynaud’s Phenomenon, esophageal dysfunction, sclerodactyly, telangiectasias). During routine clinical visits, the treating physician (rheumatologist or gastroenterologist) screened for significant symptoms of GI dysfunction. Such symptoms included refractory gastroesophageal reflux disease, early satiety, nausea/vomiting, unintentional weight loss, distension, bloating, diarrhea, and/or constipation. Patients with significant upper GI disease or lower and upper GI dysfunction symptoms who agreed to complete a whole gut transit study were included.

The Johns Hopkins Institutional Review Board approved all human patient protocols, and all individuals signed written informed consent. Paraffin sections of the adult small intestinal full-thickness gut from deidentified human tissues were obtained from the Department of Pathology at Beth Israel Deaconess Medical Center, and the Institutional Review Board approved using the deidentified human tissues.

### Clinical phenotyping of SSc

Demographic characteristics included age, sex, and race based on the standard data collection practice in the Johns Hopkins Scleroderma Center Research Registry. Clinical data such as disease duration, smoking status, SSc subtype, presence (yes/no) of telangiectasia, calcinosis, arthralgia, synovitis, and tendon friction rubs were documented by the physician at the patient’s first clinical encounter and longitudinally at 6-month intervals during routine clinical follow-up. Disease duration was defined as the interval between the first non-Raynaud’s symptom and the whole gut transit study (WGT). SSc subtype (diffuse or limited cutaneous disease) was determined based on the extent of skin involvement. GI, cardiac, muscle, and lung involvement and severity were also captured at baseline, and at the maximum severity, measures were captured longitudinally using Medsger Severity Scores^22^. Specifically, the modified Medsger GI severity score includes five categories : (a) score 0 = normal (no GI symptoms); (b) score 1 = requiring gastroesophageal reflux disease (GERD) medications (including H_2_ blocker or proton-pump inhibitor); (c) score 2 = high-dose GERD medications or antibiotics for bacterial overgrowth; (d) score 3 = episodes of pseudo-obstruction or malabsorption syndrome; and (e) score 4 = severe GI dysmotility requiring enteral or total parenteral nutrition. Sicca symptoms were defined as the presence of any of the following: dry eyes for >3 months; the sensation of sand or gravel in the eyes; the use of artificial tears three times daily; dry mouth for >3 months; swollen salivary glands; and/or the requirement of liquids to swallow due to dry mouth. The presence of overlap PBC was identified through retrospective chart review of the cohort. Participants were classified as having overlap PBC if they possessed a persistently elevated serum alkaline phosphatase with subsequent imaging and/or pathology consistent with this diagnosis. Participants without a persistently elevated alkaline phosphatase or a workup that revealed an alternative hepatobiliary pathology were classified as not having overlap PBC disease^23^.

### Whole Gut Transit (WGT) scintigraphy

All participants underwent WGT scintigraphy. Patients fasted the night before the study and were told to avoid promotility agents, antibiotics, opiates, benzodiazepines, and stool softeners three days before the study. Per protocol, patients ingested a standard amount of radiolabeled In-111 water to evaluate esophageal and liquid gastric emptying and a radiolabeled Tc-99m egg meal to evaluate solid gastric emptying. A gamma camera was used to capture anterior and posterior images of the GI tract at predetermined time points (one-half hour, 1 hour, 2 hours, 4 hours, 6 hours, 24 hours, 48 hours, and 72 hours) to assess transit throughout the entire GI tract^24^.

### Assessment of AM_2_A antibodies by enzyme-linked immunosorbent assay (ELISA)

155 serum samples from SSc-GI patients were screened for AM_2_A antibodies using the INOVA Quanta Lite M2 EP (MIT3) ELISA kit from INOVA-WERFEN (Cat. No. 704540). Per the manufacturer’s recommendations, results were interpreted as negative for <20 Units, equivocal for 20.1 – 24.9 Units, and positive for >25 Units.

### Uptake of SSc autoantibodies and immunostaining of HepG2 cells

HeLa cells were grown to 70% confluence on sterile 4 chamber slides in DMEM/High Glucose (Life Technologies) supplemented with 10% FBS (Life Technologies) at 37°C and 5% CO_2_. The cells in two chambers were cultured without human serum, while cells in the other two chambers were cultured overnight with AM_2_A-positive SSc sera diluted 1:50 (FW-2530 and FW-2340). The culture medium was then removed, and the cells were washed, followed by fixation in ice-cold 4% paraformaldehyde (10 minutes), permeabilization and blocking (performed together for 30 mins at room temperature with blocking permeabilizing buffer (BPB) comprising 5% goat serum (Life Technologies), 0.1% Triton X100 (Sigma), in PBS). The cells were then washed and incubated with antibodies against the M2 antigen (PDC-E2 protein encoded by the *Dlat* gene, Proteintech Cat. 13426-1-AP, 1:500 dilution for 1 hour at room temperature). The cells were subsequently counterstained with secondary antibodies anti-human 647 and anti-rabbit 488 (Life Technologies, 1:500 dilution for 1 hour at room temperature). Nuclei were stained with 40,6-diamidino-2-phenylindole (DAPI), and imaging was performed using Olympus confocal microscopy. Tissues were imaged under a 40X oil-immersion objective lens, with laser settings selected to ensure no overlap between fluorophores. Images were analyzed using the Fiji ImageJ software.

### Immunofluorescence staining of adult small intestinal murine longitudinal muscle – myenteric plexus (LM-MP) layer

Freshly peeled and isolated ileal LM-MP tissues from an adult male Wnt1-cre:tdTomato neural crest-specific lineage fate mapping mouse, which we have previously used to label all neural crest derivatives in the GI tract with the red fluorescent reporter tdTomato, were isolated as described^25^. Within 30 minutes of isolation, the LM-MP tissues were fixed with freshly prepared ice-cold 4% paraformaldehyde (PFA) for 5 – 10 minutes. After washing off the 4% PFA solution, the tissues were blocked and permeabilized with BPB for 30 minutes at room temperature. Next, the tissues were immunostained overnight at 16°C either with (a) paraneoplastic disease patient-derived ANNA1 antisera with demonstrated and validated specific anti-Hu immunoreactivity (1:1000), and with commercial anti-TOM20 antibody (1:500), or with (b) ANNA1 antisera (1:1000) and commercially available anti-M2 (PDC-E2/*Dlat*, Proteintech, 1:500) antibody. Secondary immunostaining was performed for 1 hour at room temperature using anti-human 647 (cyan) and anti-rabbit 488 (green) secondary antibodies (both 1:500). Nuclei were stained with DAPI, and imaging was performed and analyzed as described above.

### Immunofluorescence staining of adult small intestinal human tissue specimens

Formalin-fixed paraffin-embedded (FFPE) tissue sections of full-thickness adult human small intestine were obtained from a non-SSc patient with no known motility disease or dysfunction. Sections were immunostained with ANNA1 antisera (1:1000) and commercially available anti-M2 antibody (1:500, Proteintech). Briefly, the tissue sections were baked (60°C for 10 minutes), then blocked and permeabilized with 5% horse serum and 0.5% TritonX100 in PBS for 1 hour. The tissues were washed, then immunostained with primary antibodies overnight at 16°C. Counterstaining was performed with anti-human 647 (cyan) and anti-rabbit 488 (green) secondary antibodies (both 1:500) for 1 hour at room temperature. After washing, the sections were mounted with a Prolong Anti-fade mounting medium. Nuclei were counterstained with DAPI, and imaging was performed and analyzed as described above.

### Statistical Analysis

Clinical and demographic features were compared between AM_2_A–positive and negative patients using Chi-square or Fisher’s exact tests, Student’s t-tests, or Mann-Whitney Wilcoxon tests according to the type and distribution of the data. Spearman correlation was used to assess the correlation between two continuous variables. Regression models were used to determine whether AM_2_A antibodies were associated with GI transit. Adjusted multivariable models were developed and adjusted for potential confounders. STATA15 (STATA Corporation) and GraphPad Prism were used for statistical analyses. A p-value of < 0.05 was considered statistically significant.

## RESULTS

### Anti-M2 autoantibodies (AM_2_A) are present in a SSc cohort enriched for GI disease

The SSc cohort studied (Gastrointestinal Assessment Protocol; GAP) is enriched for patients with GI dysfunction. It is an NIH-funded cohort that was developed to study GI manifestations in SSc^7^. Among patients in this cohort, we determined that 12.9% (19/147) were AM_2_A-positive (>25 Units). Of these, none were scored as Medsger severity group 0 (“normal” GI function). Compared to patients in the cohort who were AM_2_A negative, the antibody-positive patients were also less frequently classified as having mild GI severity (Medsger group 1; gastroesophageal reflux disease, GERD) (16% vs. 28%). However, more severe physician-determined upper GI disease (Medsger 2; GERD with high doses of medications required or small bowel dilation on radiographs) was seen among AM_2_A- positive compared to negative patients (73% vs. 52%). AM_2_A positivity was less frequent among patients with Medsger Group 3 (malabsorption or intestinal pseudo-obstruction) (11% vs. 13%) and was observed in none of the 7 patients in Medsger Group 4 (require total parenteral nutrition (TPN)). No demographic or extraintestinal clinical features differed significantly between anti-AM_2_A-positive and negative patients (see Supplemental Table 1). Among the patients that were AM_2_A-positive, 15.8% (3/19) had documented overlap PBC disease; there was one patient that was classified as unknown PBC disease status due to lack of documented hepatobiliary evaluation in the setting of a persistently elevated serum alkaline phosphatase.

### AM_2_A-positive SSc patients have slower esophageal and gastric transit

We then sought to determine whether AM_2_A positivity in SSc is associated with GI transit. Using data acquired from scintigraphy-based whole gut transit studies, we determined that AM_2_A-positive patients were significantly more likely to have slower median esophageal percent emptying at 10 seconds than antibody-negative patients [69% (39, 84) vs. 82% (68, 90); p=0.03]. Furthermore, higher levels of AM_2_A antibodies correlated with worse esophageal transit (rho -0.27; p<0.01). Univariate regression analyses demonstrated an inverse relationship between AM_2_A antibody levels and percent emptying of the esophagus at 10 seconds (β-0.31, 95%CI -0.49, -0.13; p<0.01) and percent emptying of solids in the stomach at 4 hours (β-0.11, 95%CI -0.20, -0.01; p= 0.04). Interestingly, there were no associations between AM_2_A antibody levels/positivity and transit in the small bowel and colon. The above associations remained statistically significant, even after adjusting for age, race, and disease duration.

### Specific neurons in the gut wall are enriched in mitochondria and the M2 antigen

Intestinal motility is regulated by neurons and glial cells of the ENS, which are housed within the gut wall.^26^ GI dysmotility in SSc patients and the positive association between delayed GI transit and higher AM_2_A antibody levels suggest that ENS dysfunction may correlate with dysfunctional mitochondria. To examine the presence of the M2 antigen (PDC-E2 protein) in neurons of the human ENS, we immunostained sections of full-thickness small intestinal tissue obtained from a patient without SSc and any known GI motility disorder. This was performed with a commercially available anti-M2/PDC-E2 antibody and with the ANNA1 antiserum (from a patient with paraneoplastic disease and validated to contain antibodies specifically against the neuronal Hu proteins). We observed that compared to other cells in the gut wall, a subset of human enteric neurons in the myenteric ganglia showed a robust enrichment of M2 antigen (Fig 1).

**Figure 1:**
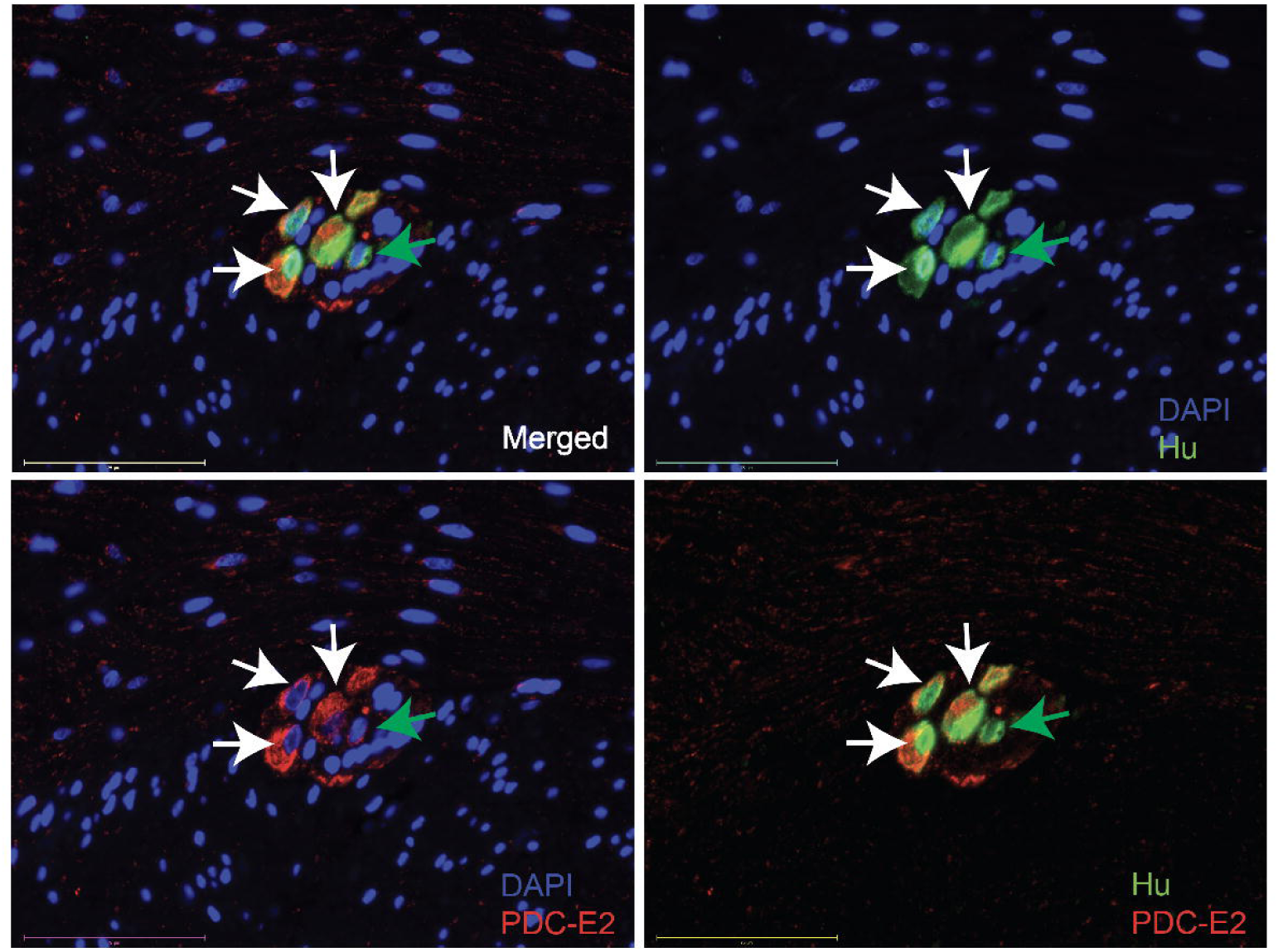
A subset of human small intestinal myenteric neurons is enriched in M2/PDC- E2 antigen. Immunostaining of section of an adult human small intestinal tissue with ANNA1 antisera containing antibodies against Hu (green) and antibodies against M2 antigen (PDC-E2; red) shows that a subset of enteric neurons (white arrows) is enriched in the PDC- E2 protein, while other neurons are not (green arrow). The representative image also shows that other cells of the gut wall are not similarly enriched in the PDC-E2 protein. Nuclei are labeled with DAPI (blue). Scale bar = 75 μm.

We recently showed that the adult mammalian ENS comprises neurons of two developmental lineages: the canonical lineage of neural crest-derived enteric neurons (NENs) and post-natal mesoderm-derived enteric neurons (MENs). These two subsets of ENS cells are molecularly distinct and contain significantly different neurochemical codes^25^. Whether NENs and MENs differ in their abundance of mitochondria and M2 antigen, thus making one of the neuronal lineages more significant in the pathophysiology of SSc patients with AM_2_A antibodies, is unknown. To address this, we immunolabeled adult murine small intestinal longitudinal muscle containing myenteric plexus (LM-MP) tissue from the neural crest lineage-fate mapping mouse model Wnt1-cre:tdTomato with neuron-specific ANNA1 antiserum and with anti-M2/PDC-E2 or pan-mitochondrial anti-TOM-20 antibodies. We observed that both M2 and TOM-20 immunoreactivity are highly enriched in the tdTomato^-^ mesoderm-derived enteric neurons (MENs) compared to other tdTomato^+^ ENS cell populations in the myenteric plexus tissue that are derived from the neural crest (Figs 2 and 3). These data show that in addition to the M2 antigen, mitochondria are enriched within the ENS cells compared to the rest of the gut wall tissue. Furthermore, within the ENS, a subset of MEN’s neurons (not the neural crest-derived cells) was enriched in mitochondria.

**Figure 2:**
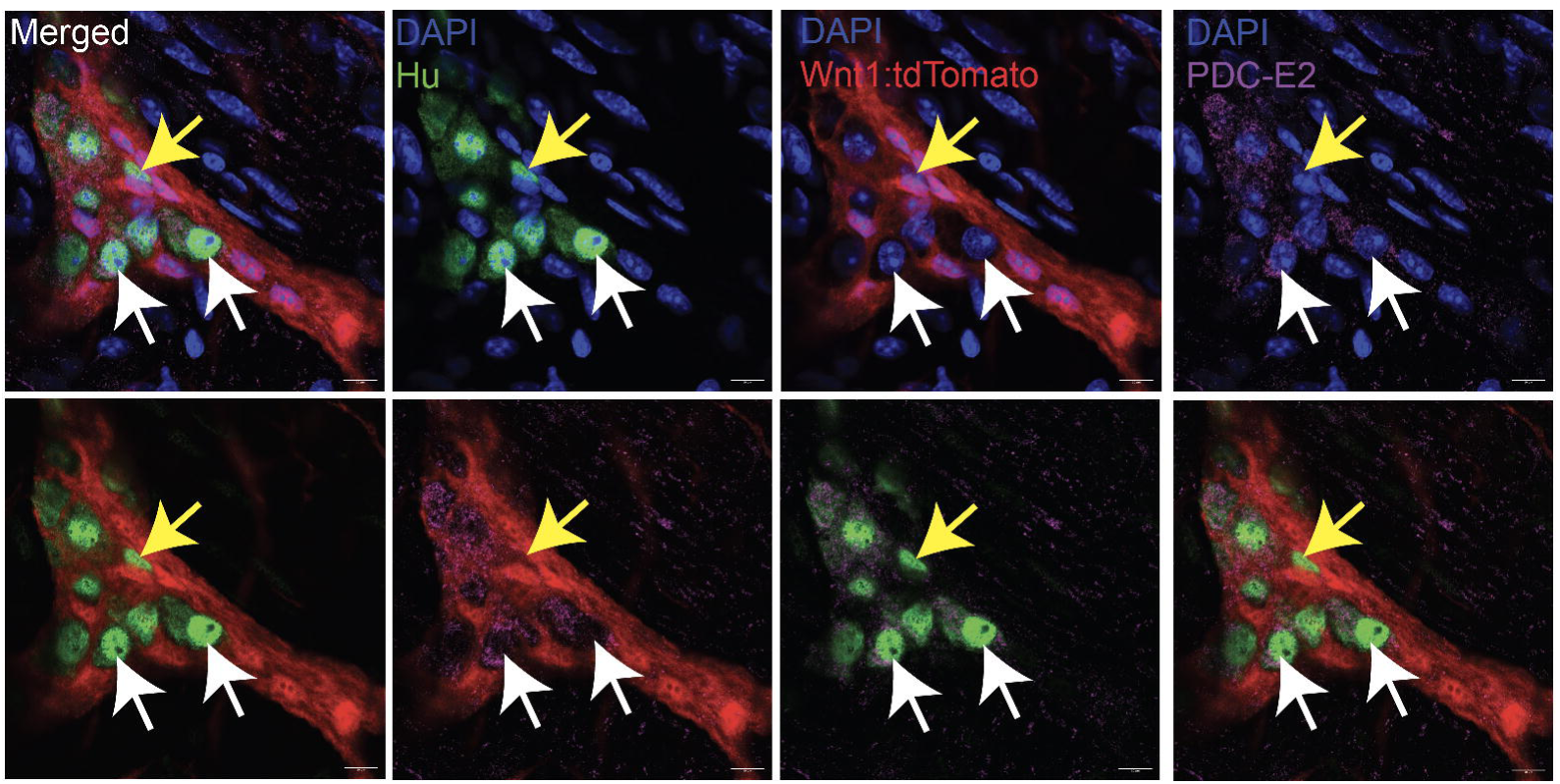
Mesoderm-derived enteric neurons (MENs) are enriched in M2/PDC-E2 antigen. Immunostaining of adult murine small intestinal longitudinal muscle - myenteric plexus (LM-MP) tissue with ANNA1 antisera containing antibodies against Hu (green) and antibodies against M2 antigen (PDC-E2; cyan). LM-MP is derived from an adult neural crest lineage-specific mouse Wnt1-cre:tdTomato, where tdTomato (red) labels neural crest-derived cells. The representative image shows that tdTomato^-^ Hu^+^ neurons (MENs, white arrows) are enriched in PDC-E2, while the neural crest-derived neurons (NENs, yellow arrow) are not. Nuclei are labeled with DAPI (blue). Scale bar = 10 μm.

**Figure 3:**
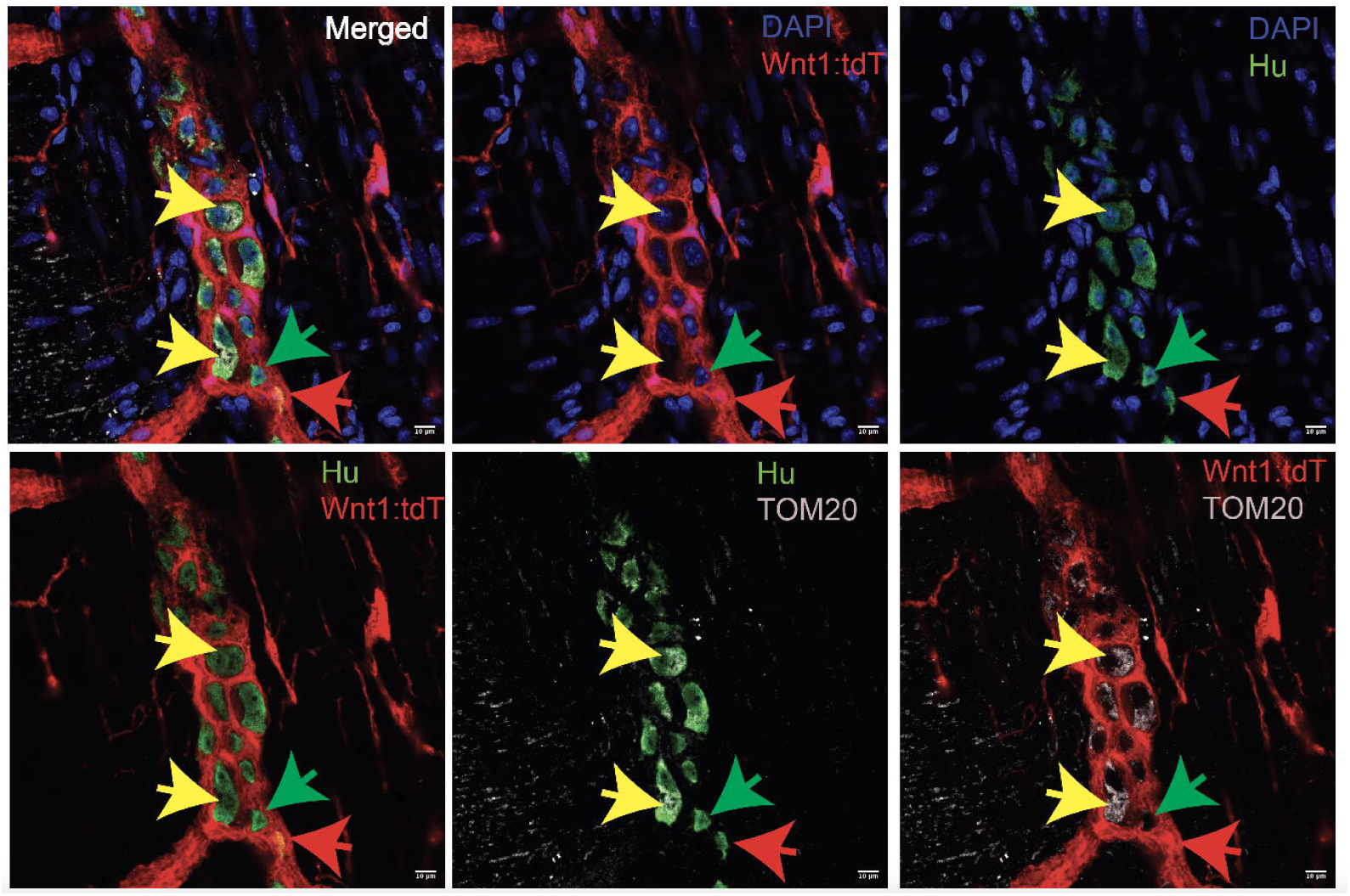
A subset of mesoderm-derived enteric neurons (MENs) are enriched in mitochondria. Immunostaining of adult murine small intestinal longitudinal muscle - myenteric plexus (LM-MP) tissue with ANNA1 antisera containing antibodies against Hu (green) and antibodies against the pan-mitochondrial antigen TOM20 (cyan). LM-MP is derived from an adult neural crest lineage-specific mouse Wnt1-cre:tdTomato, where tdTomato (red) labels neural crest-derived cells. The representative image shows that tdTomato^-^ Hu^+^ neurons (MENs, yellow arrows) are enriched in TOM20, while the neural crest-derived neurons (NENs, red arrow) and a subset of MENs (green arrow) are not. Nuclei are labeled with DAPI (blue). Scale bar = 10 μm.

### SSc autoantibodies penetrate live cells and localize to their target antigens

The presence of AM_2_A antibodies and the enrichment of M2 antigen in MENs suggest that MENs may be impacted in AM_2_A-positive SSc patients with GI dysmotility. It is currently unknown whether AM_2_A are generated due to MEN cell death and whether they regulate MEN function. Prior reports have postulated that autoantibodies against intracellular antigens may penetrate live viable cells^27^. Since the presence of MENs in the post-natal ENS is a very recent discovery^25^, conditions supporting their viability in culture have not yet been established. We therefore used cultured hepatocyte HepG2 cells, a cell line of GI origin that has been used to study mitochondrial biology^28^, to test the ability of SSc autoantibodies to penetrate relevant cells. By culturing HepG2 without patient sera, and then with two different SSc sera (with and without AM_2_A), we found that autoantibodies from both SSc sera gained access to the intracellular compartments of live viable HepG2 cells. However, only autoantibodies from the anti-AM_2_A-positive serum co-localized with the M2/PDC-E2 antigen (detected with a commercially available anti-M2 antibody). In contrast, autoantibodies from the anti-AM_2_A-negative patient did not label the cellular compartment containing the M2 antigen (Figure 4).

**Figure 4:**
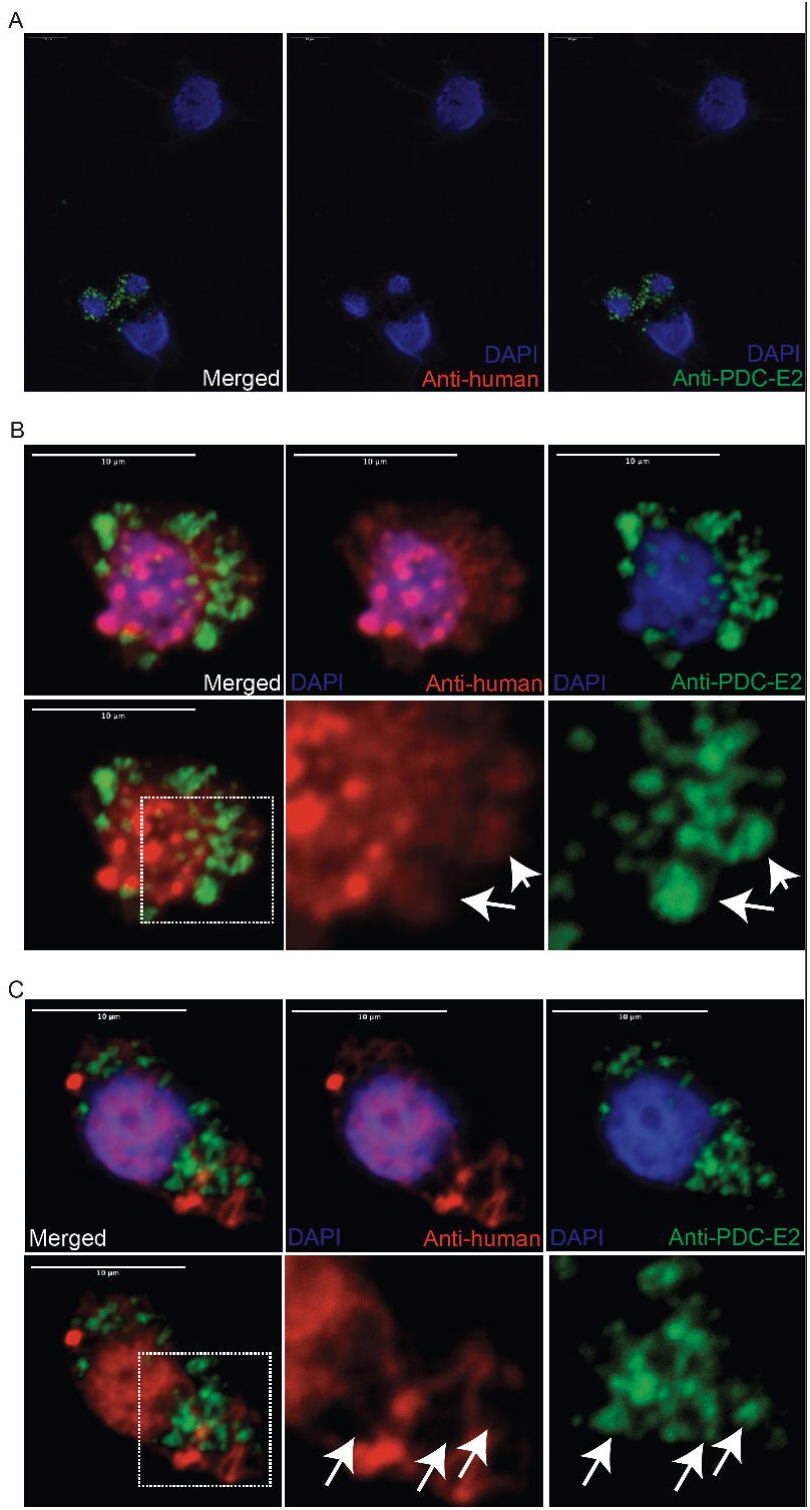
SSc autoantibodies penetrate viable cells and engage intracellular targets upon uptake. HepG2 cells when cultured with and without SSc antisera (red) can be used to test intracellular uptake. Live HepG2 cells cultured (A) without SSc antisera, and with (B) AM_2_A^+^ SSc antisera (patient FW-2530), and (C) AM_2_A^-^ SSc antisera (patient FW-2340), were subsequently fixed and immunostained with commercially available anti-M2/PDC-E2 antibody (green) and imaged to observe that SSc autoantibodies were present within cells cultured with SSc patient antisera. Furthermore, AM_2_A^+^ antisera was detected in PDC-E2- expressing mitochondria (B, white arrow), but the AM_2_A^-^ antisera labeled cellular compartments other than the PDC-E2-expressing mitochondria (C, white arrows). Nuclei are labeled with DAPI (blue). Scale bar = 10 μm.

## DISCUSSION

Our study is the first to define AM_2_A antibodies in a subset of SSc patients with well-characterized GI dysfunction and to identify a correlation between AM_2_A antibody levels and GI transit in patients with SSc. Our findings highlight a previously unappreciated link between immune responses targeted towards a specific neuron lineage in the enteric nervous system and the presence of GI disease. In addition, we show that SSc autoantibodies against mitochondrial and other antigens can penetrate viable cells, and ‘home in’ to their target antigens.

This study was performed on a large cohort of SSc patients with well-defined GI symptoms and objectively measured whole gut transit. We found that a significant proportion (∼13%) of SSc patients with GI dysfunction harbor autoantibodies against one defined mitochondrial antigen (M2). We also established that higher levels of AM_2_A antibodies are associated with slower esophageal and gastric transit in SSc patients. Using an immunostaining approach, we showed that the M2 antigen and mitochondria are enriched in enteric neurons, specifically in the recently discovered lineage of mesoderm-derived enteric neurons (MENs), relative to surrounding cell types within the ENS or the gut wall.

Prior studies have reported AMA and specifically AM_2_A in SSc patients^14, 16^, but the relevance of AM_2_A antibodies to GI dysfunction in SSc has not been described. Hoppner et al., in their SSc cohort with known cardiovascular dysfunction, estimated the prevalence of AM_2_A to be ∼10%^29^. In our cohort of SSc patients with well-characterized GI dysfunction, a similar proportion (∼13%) of patients were anti-M2-antibody positive. However, it is unclear whether the antibody titers differed between the Hoppner cohort and ours. This is important, as it was the AM_2_A antibody levels that correlated with worse GI transit (ascertained by the percent emptying of the esophagus at 10 seconds and the percent emptying of the stomach at 4 hours), suggesting a disease-associated correlation between AM_2_A antibody levels and the severity of GI dysfunction.

The M2 antigen contains two components, the E2 subunit of the Pyruvate Dehydrogenase Complex (PDC-E2) and the E2 subunit of the Branched Chain 2-Oxo Acid Dehydrogenase Complex (BCOADC-E2)^30^. Using commercially available antibodies against one of the M2 antigens (PDC-E2) as well as against a pan-mitochondrial marker (TOM20), we showed that the M2 antigen and mitochondria are abundant in murine and human ENS cells and are enriched in the lineage of mesoderm-derived enteric neurons (MENs). We previously identified the lineage of MENs distinct from neural crest-derived enteric neurons (NENs). Enrichment of mitochondria and M2 antigens in neurons of this lineage suggests that the dysmotility observed in the AM_2_A-positive SSc-GI patients negatively impacts this lineage of neurons. The exact nature of the mechanisms through which MENs may be involved in the AM_2_A-mediated pathogenesis of GI dysmotility remains to be elucidated and will require dedicated mechanistic studies using animal models.

Our study has several strengths. First, we established that AM_2_A are present in a significant proportion of SSc patients and correlate inversely with GI transit. Our finding that of all the cells in the gut wall, ENS neurons are enriched in mitochondria and mitochondrial antigens, provides a possible rationale for why patients with pathologies that target or impact mitochondria (e.g., presence of AM_2_A) overwhelmingly impact the ENS to drive GI dysfunction. Furthermore, our study extends this by showing that only a subset of ENS neurons – those that we recently discovered are developmentally derived from mesoderm^25^ – are specifically enriched in mitochondria relative to the other cell populations. These data pinpoint MENs as the lineage most dependent on and susceptible to mitochondrial function in the gut wall and suggest that mitochondrial dysfunction overwhelmingly impacts MENs to drive GI dysfunction. The presence of AM_2_A and GI dysfunction in SSc patients suggests that MENs are impacted in the pathobiology of disease but leaves the question of how these autoantibodies impact the mitochondria-laden MENs. Limitations of our study include the fact that the SSc patient cohort is limited to a relatively small number of patients with intestinal dysmotility; hence, the relevance of AM_2_A to small bowel dysmotility cannot be fully understood. Our technical limitation of not yet being able to culture MENs restricts us to performing the AM_2_A penetration studies on other cell types instead of MENs. Finally, our current study does not involve examining the impact of AM_2_A on *in vivo* gut biology in a model organism. Future studies focused on the ability of AM_2_A to drive GI dysfunction in a model system are warranted.

That SSc autoantibodies can penetrate live cells and transit intracellularly to their cognate antigens is a noteworthy finding. It raises the tantalizing possibility of subsequent functional consequences that might include corrupting the functioning of important organelles and cellular behaviors to impact tissue and organ functions negatively. Our study highlights the importance of studying the incidence of dysmotility in various GI organs in carefully phenotyped AM_2_A-positive SSc patients to better understand the spectrum of GI dysmotility. Knowledge gained from such studies will provide important insights into the possible role of AM_2_A antibodies in the pathophysiology of GI dysmotility in SSc and other autoimmune conditions.

## Data Availability

All data produced in the present work are contained in the manuscript

## ACKNOWLEDGMENTS

This work was supported by funding from NIAMS R01 AR081382-01A1 (ZM), the Rheumatology Research Foundation and Scleroderma Research Foundation (ZM), NIA R0AG166768, DIACOMP pilot funding, and pilot and feasibility grant from Harvard Digestive Disease Core (all three to SK), R01 AR073208 (LCR), and the Donald B. and Dorothy L. Stabler Foundation. The Rheumatic Diseases Research Core Center, where the ELISA assays were performed, is supported by NIH P30-AR070254. We would also like to acknowledge the contribution of the JHU scleroderma center, physicians, support staff and patients.

